# Fidelity-Derived Quantum Dissimilarity-Enhanced *k*-Nearest Neighbor Algorithm for Arterial Hypertension Prediction

**DOI:** 10.64898/2026.06.08.26355139

**Authors:** Antonia E. Tampakaki, Georgios D. Barmparis, Eleni Angelaki, Maria E. Marketou, Giorgos P. Tsironis

## Abstract

We present a quantum-enhanced version of the classic *k*-Nearest Neighbors (*k*NN) classification algorithm, applied to the prediction of arterial hypertension. The traditional Euclidean distance metric of the *k*NN algorithm is replaced with a Fidelity-derived quantum dissimilarity measure to evaluate the similarity between data samples. We map classical real-world clinical and ECG-derived data features into quantum states via the Dense-Angle Encoding, which efficiently utilizes parameterized rotation gates to pack multiple features into minimal qubits while maintaining pure states. We evaluate the performance of the dissimilarity measure using both the noiseless state vector Simulator and the IBM Qiskit Estimator primitives. The quantum circuit demonstrates robust predictive capabilities comparable to the classical model. While it does not claim computational supremacy over the classical baseline, the framework proves that fidelity-based similarity is a physically meaningful and efficient approach for hybrid quantum-classical classification.

## I. INTRODUCTION

Quantum machine learning (QML) has emerged as a rapidly developing interdisciplinary field that combines quantum information theory with modern machine learning (ML) methods. The growing interest in QML is closely connected to the development of noisy intermediate-scale quantum (NISQ) devices [1], which provide experimentally accessible platforms for hybrid quantum-classical algorithms that overcome current hardware limitations of Quantum Computers (QCs). By nature, QCs cannot operate without interacting with the environment. This interaction, known as quantum noise, introduces errors in quantum systems through decoherence, gate errors, and limited qubit counts [2]. Although these errors can be mitigated by quantum error correction protocols [3, 4], overcoming them remains a critical hurdle for scaling quantum computers to practical QML applications. Recent research in the NISQ era has focused on shallow quantum circuits, variational quantum algorithms [5], quantum kernels, and Fidelity-based learning methods [6] that can operate under realistic hardware constraints [7].

In order to apply a QML classification algorithm to classical data, the data must be converted to quantum data. Converting classical data into a quantum format involves mapping it onto quantum bits (qubits), the fundamental units of information in quantum computers. This process is known as encoding, and several techniques are available, including basis, angle, and amplitude encoding [8]. Sometimes the input process can be much more expensive and time-consuming than the quantum algorithm itself [9]. Utilizing angle encoding, Ovalle-Magallanes et al. [10] concluded that the angle encoding of the data has a significant impact on the capabilities of convolutional neural networks. Salinas et al. [11] proved that data encoding in a single qubit within the angle parameters of the rotation gates was an fulfilling method to achieve classification. Deluca et al. [12] compared different methods of encoding, including angle and amplitude encoding, to conclude that each method significant influences on the final classification result. Additionally, Sein et al. [13] chose angle encoding method because of its simplicity compared to the other quantum encoding methods to evaluate different model types for classification.

Among supervised learning methods, the k-Nearest Neighbors (kNN) [14] algorithm remains one of the simplest and most interpretable model-free classification techniques. Conventional kNN relies on geometric distance measures, typically the Euclidean metric, to quantify similarity between data samples. Several quantum variants of the *k*NN algorithm have been proposed. Zardini et al. [15] used the amplitude encoding and the Euclidean distance estimation. The amplitude encoding was also used by Estrada et al. [16], who used non-binary numerical data and compared quantum states using the Hamming distance. Quezada et al. [17] compare the classical kNN with the Quantum kNN by working on the depth of the circuit and all the parameters that come with it. Schuld et al. [18] used the Hamming distance and the assumption that the closest vector also has the class information of the test point.

In this work, we apply a quantum-enhanced kNN classifier in which Euclidean distance is replaced by a Fidelity-derived quantum dissimilarity measure. Fidelity [6, 19] between density matrix operators measures the overlap between quantum states and provides a physically meaningful measure of their distinguishability. For pure quantum states, fidelity reduces to the square of the overlap between the quantum states. This interpretation makes fidelity particularly suitable for QML applications involving state preparation and quantum feature embeddings. The proposed framework encodes real-world clinical and ECG-derived data for predicting arterial hypertension, as described in Ref. [20], into quantum states using parametrized rotation gates, especially the dense encoding. In contrast to the SWAP test, which requires an ancilla qubit and a deeper circuit structure, the proposed framework evaluates state similarity with only two qubits to classify individuals as normotensive or hypertensive. The resulting dissimilarity function is directly related to quantum-state fidelity and can be extended to higher-dimensional state representations via tensor-product decompositions. The architecture is specifically designed for NISQ compatibility by minimizing circuit depth, gate complexity, and qubit overhead while preserving the fidelity-based similarity structure required for classification. The proposed framework is implemented using IBM Qiskit [21] primitives, including the state vector Simulator for noiseless quantum-state evolution and the Estimator primitive for expectation-value evaluation under realistic execution conditions. The quantum classifier is compared against a classical kNN baseline using standard classification metrics. Our objective is not to claim computational quantum advantage, but rather to investigate whether fidelity-based quantum similarity measures can provide a robust and physically meaningful framework for hybrid quantum-classical classification in biomedical applications.

The paper is organized as follows. In Section II, we present the methodology used for processing the data, developing the classification algorithm, and defining the evaluation metrics. In Section III, we present the results of the quantum simulator and the quantum estimator. Finally, we discuss implications and future directions in Section IV.

## II. METHODS

### A. Data preparation

We studied 1091 individuals aged 30–80 years of both sexes, with and without essential arterial hypertension, without cardiovascular diseases (CVD), as examined by two Cardiology centres from November 2019 to October 2021, each one described by the 6 most important anthropometric and ECG-derived features as found in Ref. [20], i.e., age, sex, body fat (BF), R wave amplitude in aVL (R in aVL), body mass index adjusted by Cornell criteria [BMI multiplied by RaVL + SV_3_] (BMI-adj Cornell), and body mass index modified by Sokolow-Lyon voltage [BMI divided by SV_1_ + RV_5_], (BMI-modified Sokolow-Lyon). We preprocessed the data by standardized them as in Ref. [20]. In quantum machine learning, all features are represented as angles; thus, we need to convert classical features into angle representations to encode them into quantum states. We apply a min-max scaler as implemented in Scikit-Learn [22] in the interval [0, *π*] (for the features encoded as Y-axis rotations) and [0, 2*π*] (for the features encoded as Z-axis rotations. See Section II B), to convert the features into angular variables for preparing the quantum states of our quantum circuit. In Fig. 1, we present the pairwise scatterplot matrix of the scaled features for the hypertensive and normotensive individuals.

**FIG. 1.**
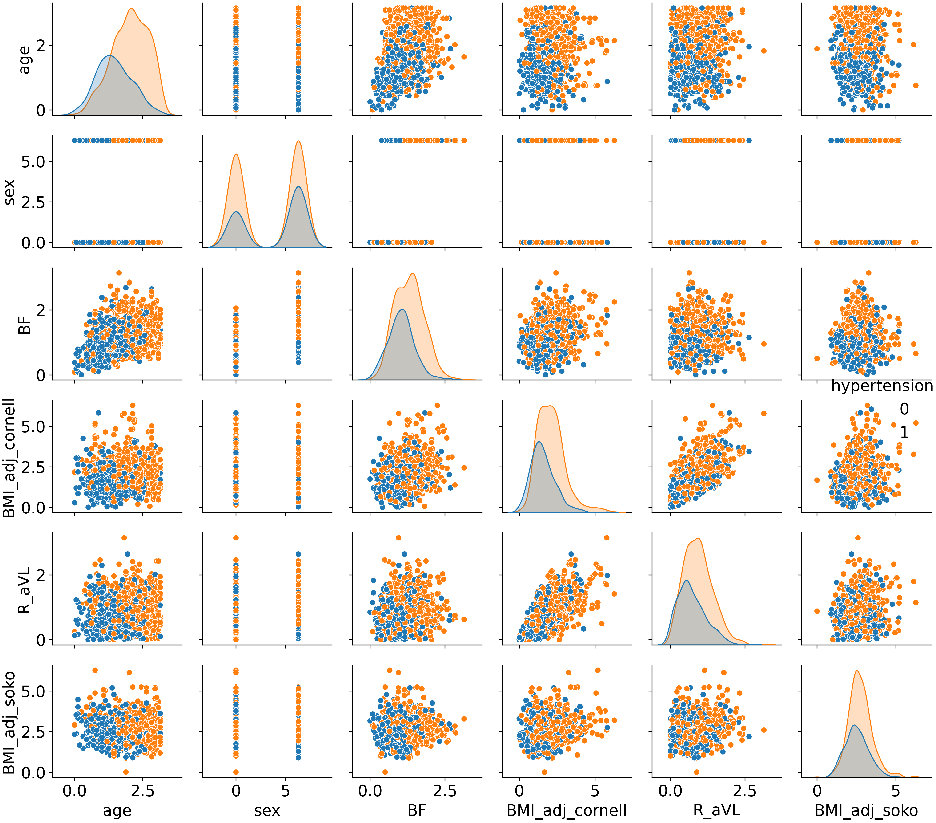
Pair plot comparison of the scaled features.

### B. Quantum Encoding

There are several robust methods to encode classical data into quantum states [23]. In this work, we apply the Dense-Angle encoding (DAE), which combines the angle encoding (single Y-axis rotation by a *θ* angle of the |0⟩ state of a qubit), and the phase encoding (single Z-axis rotation by a *ϕ* angle on the Hadamard prepared |0⟩ state), allowing two features to be encoded in a single qubit simultaneously. DAE is highly efficient because it packs 2*N* features into just *N* qubits. Under DAE, a data point described by two angle features *θ*∈ [0, *π*], and *ϕ*∈ [0, 2*π*) is encoded into a single qubit quantum state, *ψ*, as:

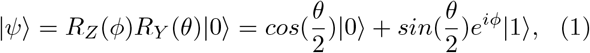

and can be visualized as a point on the surface of a Bloch Sphere, as shown in Fig. 2. To encode more features under the DAE, we need to generalize from a single-qubit state to an *n*-qubit system utilizing the Kronecker product. An *n*-qubit state can encode an even number of *N* classical features as:

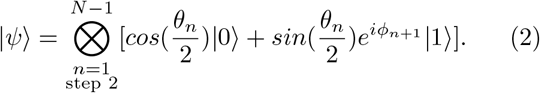

**FIG. 2.**
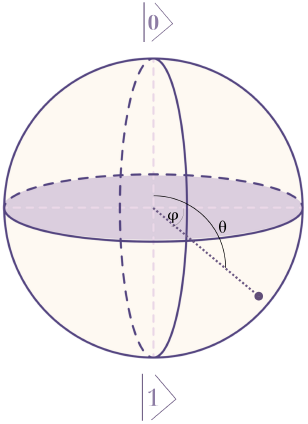
Representation of a quantum state, *ψ*, on a Bloch Sphere.

For an odd number of features, we add a zero phase to the last qubit, without affecting the final state vector. This encoding process can be performed via a Parameterized Quantum Circuit (PQC) [24] mathematically expressed as:

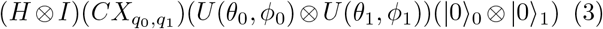

where the two qubits 0, 1 are prepared in the |0⟩ state, *U* (*θ*_*i*_, *ϕ*_*i*_) = *R*_*Z*_(*ϕ*_*i*_)*R*_*Y*_ (*θ*_*i*_), with *i* = 0, 1, represents a rotation operator applying an *R*_*Y*_ (*θ*_*i*_) and an *R*_*Z*_(*ϕ*_*i*_) rotation on each qubit, 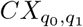is the CNOT gate, and H and I are the Hadamard and the Identity gates. The circuit is shown in Fig. 3.

**FIG. 3.**
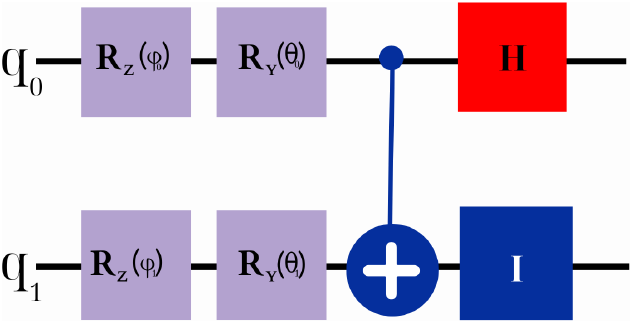
Visualization of the quantum circuit defined in Eq. (3).

Utilizing Eq. (3), the six classical features representing each individual in our dataset are encoded into three pairs of single-qubit states grouped according to feature type as: 1^*st*^ qubit - age, and sex, 2^*nd*^ qubit - body fat, BMI adjusted Cornell, and 3^*rd*^ - R in aVL, BMI-modified Sokolow-Lyon. Finally, each data point is represented by a tensor product of 3 Bloch Spheres (BS) as shown in Fig. 4, where the first BS encodes the first two features, the second BS the next two, and the third one the last two features.

**FIG. 4.**
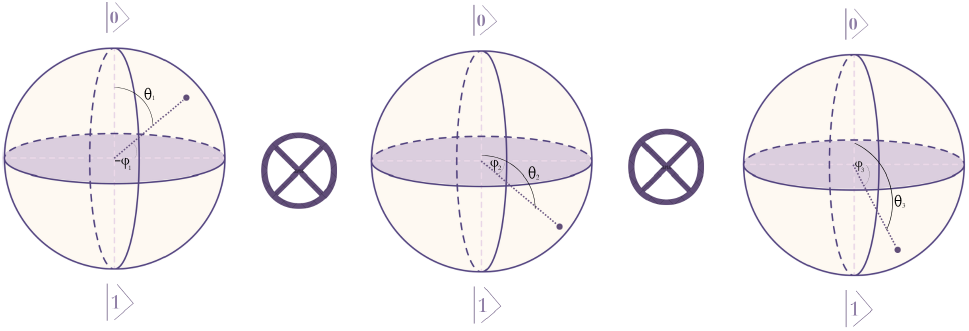
The quantum representation of a classical data point. Each data point is encoded into a quantum state as the tensor product of 3 Bloch Spheres, each one encoding two classical features.

An advantage of the DAE is that the encoded states are pure. This is because the encoding is done using unitary operations such as rotation gates (*R*_*Y*_) and phase shift gates (*R*_*Z*_), acting on a known initial state |0⟩ ^⊗*n*^ of *n* qubits. This advantage allows simplifying the Fidelity to the Quantum Dissimilarity as the metric for identifying the neighbors of a given sample in the *k*NN algorithm.

### C. Dissimilarity enhanced *k*-Nearest Neighbors

The *k*-Nearest Neighbors algorithm is considered one of the simplest supervised ML algorithms. The algorithm classifies a given sample (test point) by identifying its *k*-closest data points (neighbors) in a training set using a distance metric and predicting the majority class of those neighbors.

In the proposed *k*NN algorithm, the Euclidean distance is replaced by a Fidelity-derived quantum dissimilarity measure. Fidelity [6], *F* (*ρ*_1_, *ρ*_2_), is a measure of how similar two density matrices, *ρ*_1_ = |*ψ*_1_⟩ ⟨*ψ*_1_| and *ρ*_2_ = |*ψ*_2_⟩ ⟨*ψ*_2_|, of generally mixed quantum states is. When |*ψ*_1_⟩ and |*ψ*_2_⟩ are pure states, fidelity can be calculated as (see Appendix A):

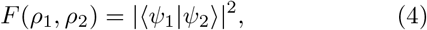

and the dissimilarity, *d*_*q*_, as:

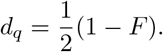

Here, we use the state vector formalism by measuring on the |11⟩ state to calculate the dissimilarity, Δ(*ψ*^*A*^, *ψ*^*B*^), between two quantum states 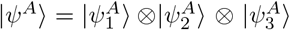, and 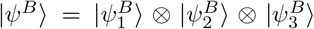 as (see Appendix B):

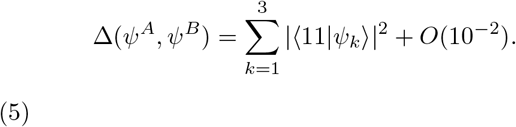

For a quantum state encoded by three single-qubit states, Eq. (II C) reduces to:

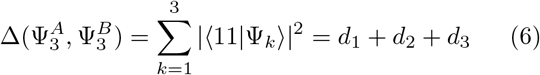

where *d*_1_, *d*_2_, and *d*_3_ are the single-qubit dissimilarities of each one of the three qubits describing the quantum states.

### D. Qiskit Estimator

The dissimilarity of a 2-qubit state |*ψ*⟩ = |*ψ*_1_⟩ ⊗ |*ψ*_2_⟩ can be estimated by mapping it to an expectation value via a Qiskit Estimator (QE). An Estimator is a Qiskit primitive, a standardized mathematical routine used to compute expectation values of observables and represents the final simulation step before execution on quantum hardware. For a 2-qubit state expressed as: |*ψ*⟩ = *α*|00⟩+ *β*|01⟩ + *γ*|10⟩ + *δ*|11⟩, where *α, β, γ*, and *δ* are in general complex numbers, the dissimilarity can be calculated as the expectation value of the density matrix, *ρ* = |*ψ*⟩ ⟨*ψ*| in the 11 state, i.e., *d*_*q*_ = ⟨11 |*ρ*| 11⟩ . Thus, by using the Pauli matrices **X, Y**, and **Z**,

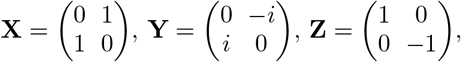

and the Identity matrix, **I**:

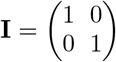

dissimilarity can be expressed as (keeping only terms that survive the |11⟩ projection):

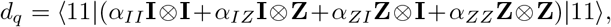

where *α*_*II*_, *α*_*IZ*_, *α*_*ZI*_, and *α*_*ZZ*_ ∈ ℝ, are the corresponding Pauli coefficients.

By expanding the operators, we conclude that the dissimilarity can be calculated in terms of the Pauli coefficients as:

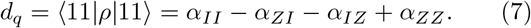

In Qiskit, Eq. (7), is evaluated as: 1

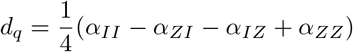

where 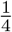 is a normalization constant.

## III. RESULTS

The performance of a *k*NN algorithm depends strongly on the available data. Thus, we use all the available data as the training set, and we apply 5-fold cross-validation by optimizing F1-score to estimate the best number of neighbors, *k* (between all odd values in the range [1, 15]) and to evaluate the performance of the *k*NN algorithm in terms of accuracy and F1-score. The 5-fold CV is applied via Scikit-Learn pipelines to ensure correct scaling in each fold. This evaluation uses both the Euclidean distance (classical) and the dissimilarity metric (quantum) calculated from the state vector and the Estimator methods. In Table I, we present the best number of neighbors, *k*, along with the mean value and the standard deviation of the accuracy and the F1-score for each method. We found that the Euclidean distance performs better when labeling data by their 11 closest neighbors, while the state vector and the Estimator perform better by their 5 neighbors. The classical *k*NN algorithm achieves an accuracy of (75.5 ± 3.0)%, and an F1-score (82.1 ± 2.4)% while the Simulator and the Estimator have slightly lower accuracy and F1-score at (72.5 ± 1.8)% and (79.2 ± 3.0)%, respectively.

**TABLE I.** Performance of the *k*NN algorithm using Euclidean and quantum dissimilarity distance on the training set using 5-fold cross-validation.

| Method | Best $k$ | Accuracy (%) | F1 score (%) |
| --- | --- | --- | --- |
| Euclidean distance | 11 | $75.5 \pm 3.0$ | $82.1 \pm 2.4$ |
| State vector | 5 | $72.5 \pm 1.8$ | $79.2 \pm 3.0$ |
| Estimator | 5 | $72.5 \pm 1.8$ | $79.2 \pm 3.0$ |

In Fig. 5, we present the normalized confusion matrices for these evaluations. We observe that the classic *k*NN algorithm predicts fewer normotensive individuals, 61.6% vs 67.7% of the state vector and Estimator implementations, while it predicts more hypertensive individuals, 89.7% vs 88.3%, respectively. In addition, the classic *k*NN classifier yields more false positives and fewer false negatives.

**FIG. 5.**
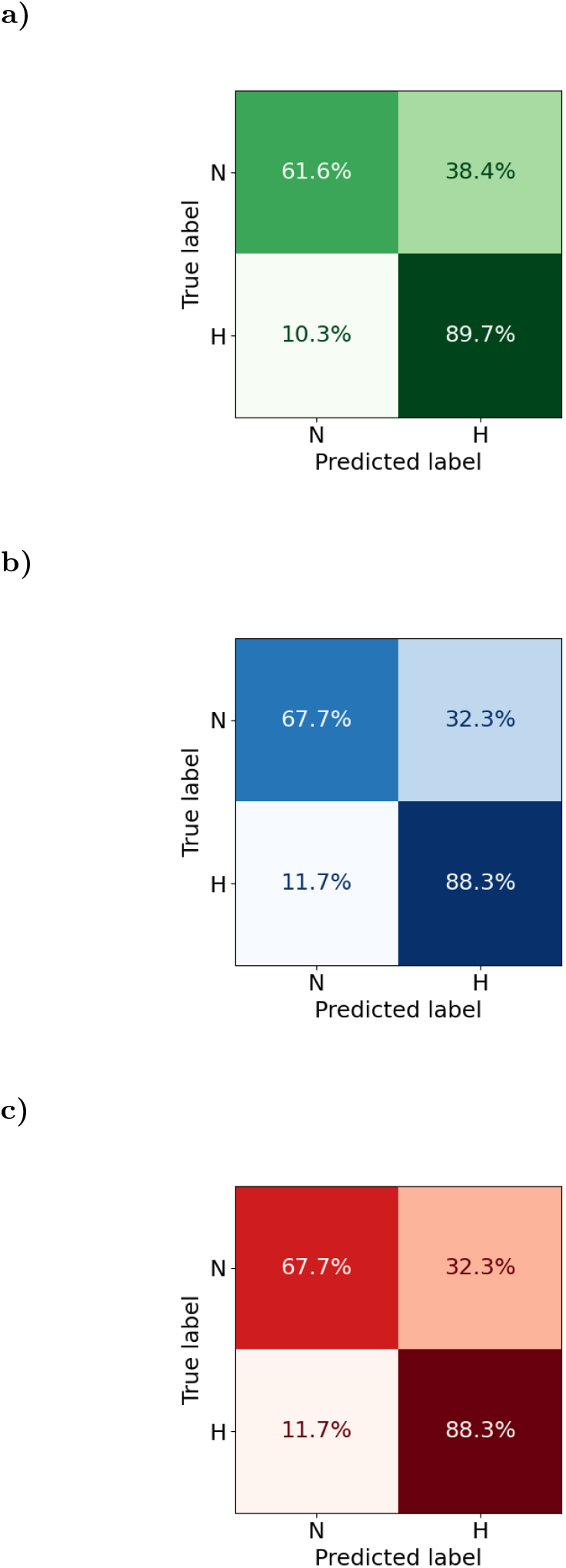
The normalized confusion matrix: a) Classical *k*NN, b) state vector, and c) Estimator. N represents the Normotensive and H the hypertensive individuals.

## IV. DISCUSSION

Quantum Machine Learning (QML) holds transformative potential for medical applications, primarily by leveraging quantum phenomena such as superposition and entanglement to enhance model performance. In this work, we demonstrated that even a fundamentally simple machine learning algorithm, such as *k*-Nearest Neighbor can be effectively adapted using a quantum comparison process, performing competitively with its classical counterpart. By utilizing Dense-Angle Encoding, we successfully minimized qubit overhead, mapping six classical clinical features into a highly efficient three single-qubit state. This ensures the architecture is specifically compatible with Noisy Intermediate-Scale Quantum (NISQ) devices, where minimizing circuit depth and gate complexity is essential. Our results show that using the quantum dissimilarity metric with the Qiskit Estimator yields an F1-score of 79.2%, which is reasonably comparable to the 82.1% obtained by the classical Euclidean model. Although the classical model maintained a slightly higher overall accuracy, the quantum adaptation remained competitive.

It is important to emphasize that the primary objective of this research was not to demonstrate immediate computational quantum advantage over classical machine learning. Achieving true quantum advantage remains a complex, long-term milestone for the scientific community. As noted in the literature, demonstrating good performance on a specific dataset does not in itself confirm a strict quantum advantage.

In conclusion, Fidelity-derived quantum dissimilarity is a reliable, physically meaningful measure for classification. Future research can extend this resulting dissimilarity function to higher-dimensional state representations via tensor-product decompositions, paving the way for more complex biomedical diagnostic tools in the QML landscape.

## Data Availability

All data produced in the present study are available upon reasonable request to the authors

## ACKNOWLEDGMENTS

This work relates to the Department of Navy Grant N629092412119 issued by the Office of Naval Research Global, USA.

## Appendix A: State vector - Measurement on |11⟩

Applying Eq(3) on two pure states |*ψ*_0_⟩ and |*ψ*_1_⟩ as produces by the DAE formalism:

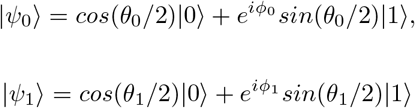

we obtain:

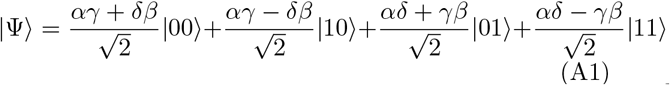

here, 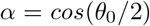, *γ* = *cos*(*θ*_1_*/*2), and 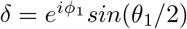, with |*α*|^2^ +|*β*|^2^ = 1, |*γ*|^2^ +|*δ*|^2^ = 1, and *α, γ* ∈ **R**.

Measuring the probability of |Ψ⟩ being in |11⟩ is equal to:

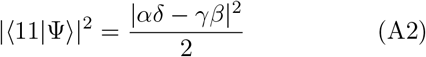

Expanding Eq. (A2) and by adding and subtracting the terms |*α*|^2^|*γ*|^2^ and |*β*|^2^|*δ*|^2^, we obtain:

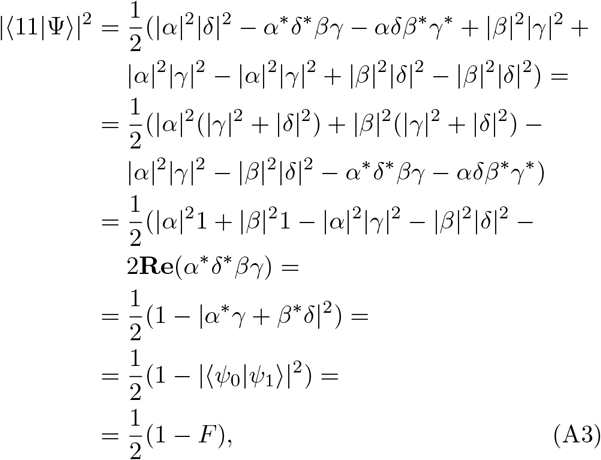

where *F* describes Fidelity.

## Appendix B: Dissimilarity for *n*-qubit states

By using the Multiplicativity property [6], the Fidelity, *F* (*ρ*^*A*^, *ρ*^*B*^), between the density matrices of the two 2-qubit pure states |*ψ*^*A*^⟩ and |*ψ*^*B*^⟩ is given as:

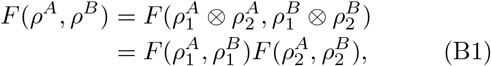

and the dissimilarity, Δ(Ψ^*A*^, Ψ^*B*^), as:

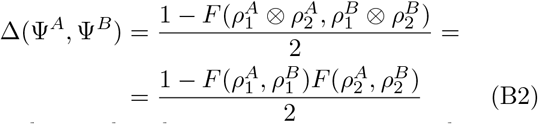

Generalizing the above expressions to *n*-qubit pure states:

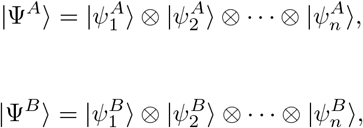

the Fidelity between Ψ_*A*_, Ψ_*B*_ is given by:

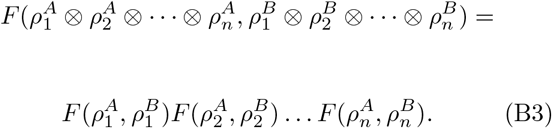

Thus the dissimilarity between the Ψ^*A*^, Ψ^*B*^ is written as:

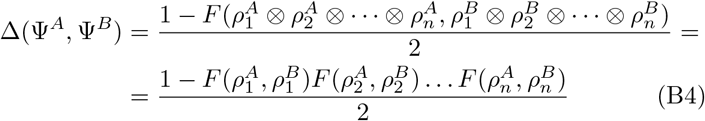

Given that each of these density matrices represents a pure state, we can replace the fidelity of each single-qubit state with its single-qubit dissimilarity, 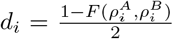 in Eq. (B3), and obtain the total dissimilarity as:

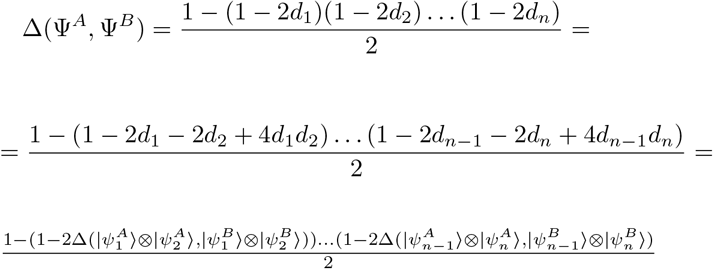

which leads to:

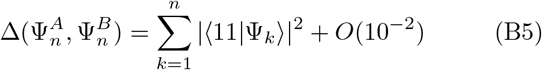

where 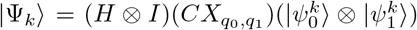 as defined by the quantum circuit in Eq. (3) under the DAE.

